# STIMULATE-ICP-CAREINEQUAL - Defining usual care and examining inequalities in Long Covid support: protocol for a mixed-methods study (part of STIMULATE-ICP: Symptoms, Trajectory, Inequalities and Management: Understanding Long-COVID to Address and Transform Existing Integrated Care Pathways)

**DOI:** 10.1101/2022.05.06.22274658

**Authors:** Mel Ramasawmy, Yi Mu, Donna Clutterbuck, Marija Pantelic, Gregory Y. H. Lip, Christina van der Feltz-Cornelis, Dan Wootton, Nefyn H Williams, Hugh Montgomery, Rita Mallinson Cookson, Emily Attree, Mark Gabbay, Melissa Heightman, Nisreen A Alwan, Amitava Banerjee, Paula Lorgelly, the STIMULATE-ICP consortium

## Abstract

**Introduction:** Individuals with Long Covid represent a new and growing patient population. In England, fewer than 90 Long Covid clinics deliver assessment and treatment informed by NICE guidelines. However, a paucity of clinical trials or longitudinal cohort studies means that the epidemiology, clinical trajectory, healthcare utilisation and effectiveness of current Long Covid care are poorly documented, and that neither evidence-based treatments nor rehabilitation strategies exist. In addition, and in part due to pre-pandemic health inequalities, access to referral and care varies, and patient experience of the Long Covid care pathways can be poor.

In a mixed methods study, we therefore aim to: (1) describe the usual healthcare, outcomes and resource utilisation of individuals with Long Covid; (2) assess the extent of inequalities in access to Long Covid care, and specifically to understand Long Covid patients’ experiences of stigma and discrimination.

**Methods and analysis:** A mixed methods study will address our aims. Qualitative data collection from patients and health professionals will be achieved through surveys, interviews and focus group discussions, to understand their experience and document the function of clinics. A patient cohort study will provide an understanding of outcomes and costs of care. Accessible data will be further analysed to understand the nature of Long Covid, and the care received.

**Ethics and dissemination:** Ethical approval was obtained from South Central - Berkshire Research Ethics Committee (reference 303958). The dissemination plan will be decided by the patient and public involvement and engagement (PPIE) group members and study Co-Is, but will target 1) policy makers, and those responsible for commissioning and delivering Long Covid services, 2) patients and the public, and 3) academics.

## Introduction

Covid-19 disease results from acute SARS-CoV-2 viral infection. Whilst many patients may remain asymptomatic, or suffer self-limiting disease, symptoms persist in some for an extended period. Long Covid “occurs in individuals with a history of probable or confirmed SARS CoV-2 infection, usually 3 months from the onset of COVID-19 with symptoms, and that last for at least 2 months and cannot be explained by an alternative diagnosis” [1]. It spans multiple healthcare challenges, including physical and psychological symptoms and functional impairment that may be episodic for some and unrelenting for others. It has been estimated that up to 12% of non-hospitalised individuals with COVID-19 may have persistent symptoms for more than 3 months [2]. The nature of the illness, together with its prevalence [3], suggests that Long Covid results in a significant individual, health care system, and societal burden [4] – [9] which may challenge the delivery of effective, high-quality and sustainable care.

Despite a growing patient population, there is no evidence-based treatment or rehabilitation intervention for Long Covid. Further, non-hospitalised patients and assessments of clinical care pathways have been under-represented in the few Long Covid clinical trials or longitudinal cohort studies so far reported [10]. The epidemiology, clinical trajectory, healthcare utilisation and effectiveness of current care for Long Covid are thus poorly documented [11],[12]. Currently in England, there are fewer than 90 specialist Long Covid clinics in which assessment and treatment is informed by NICE guidelines. However, there is evidence that access to such clinics and related care pathways, the nature of those pathways, and patient experience, varies [13]. Research is thus required to inform diagnosis, care, public health strategies, policy planning, resource allocation and budgeting. It is likewise essential to define the usual care pathway in Long Covid clinics, and to understand patient presentation, and the efficacy and cost of care.

It is important to understand how access to Long Covid services varies, and the barriers to such access and thus to diagnosis and subsequent care. Early evidence suggests that systemic inequalities in referral, assessment and rehabilitation may exist, exacerbated in part by stigma towards specific Long Covid population subgroups, based on demographic or socioeconomic characteristics [7]. It is thus important to document and understand these inequalities and how they relate to pre-pandemic health inequalities, in the new contexts of NHS pressures causing additional pressures on access, such that all patients have access to appropriate care at the right time.

Funded by the National Institute for Health Research (NIHR), the STIMULATE-ICP (Symptoms, Trajectory, Inequalities and Management: Understanding Long-COVID to Address and Transform Existing Integrated Care Pathways) project combines clinical and epidemiological studies; a large randomised controlled trial (RCT) exploring the benefit of an integrated care pathway (ICP) for Long Covid; and mixed methods studies exploring inequalities of care and transferability of the ICP to other long-term conditions (LTCs).

This protocol is for the sub-study, STIMULATE-ICP-CAREINEQUAL, which will provide much needed evidence regarding the nature of usual care, the outcomes such care delivers, costs of such care, and inequalities which currently exist with respect to accessing care. Findings will help inform development and delivery of effective and equitable case identification, support management and treatment for Long Covid.

## Aims and objectives

The aim of STIMULATE-ICP-CAREINEQUAL is to understand current Long Covid care and who is presenting at Long Covid clinics.

Specific objectives are:

1. To describe usual care, health outcomes and healthcare utilisation of individuals with Long Covid. This will include describing:
  i. Current usual care of Long Covid at each trial site involved in the STIMULATE-ICP RCT
  ii. The burden of illness at presentation to Long Covid clinics
  iii. Patient outcomes from current care pathways
  iv. Resource utilisation and the cost of current care at the patient and service level
2. To assess the extent of inequalities in access to Long Covid care. This will include describing:
  i. Whether patients who are currently accessing Long Covid services are representative of the overall population of people living with Long Covid, and if there are inequalities in access to care and outcomes.
  ii. The barriers and facilitators to accessing Long Covid diagnosis, care and support
  iii. The nature and extent of stigma and discrimination experienced by Long Covid patients.

## Materials and Methods

In order to collect and analyse data in the most pragmatic and effective and timely manner, we opted for a mixed methods approach to address these research questions. An overview of the research components, data collection methods and analyses are set out in Figure 1. Patients and health professionals will be interviewed to understand their experience and document how the clinics function, while patient data from Long Covid clinics will be explored to understand the nature of Long Covid and the care received. A follow up survey of patients will provide an understanding of the outcomes of care and the cost.

**Figure 1:**
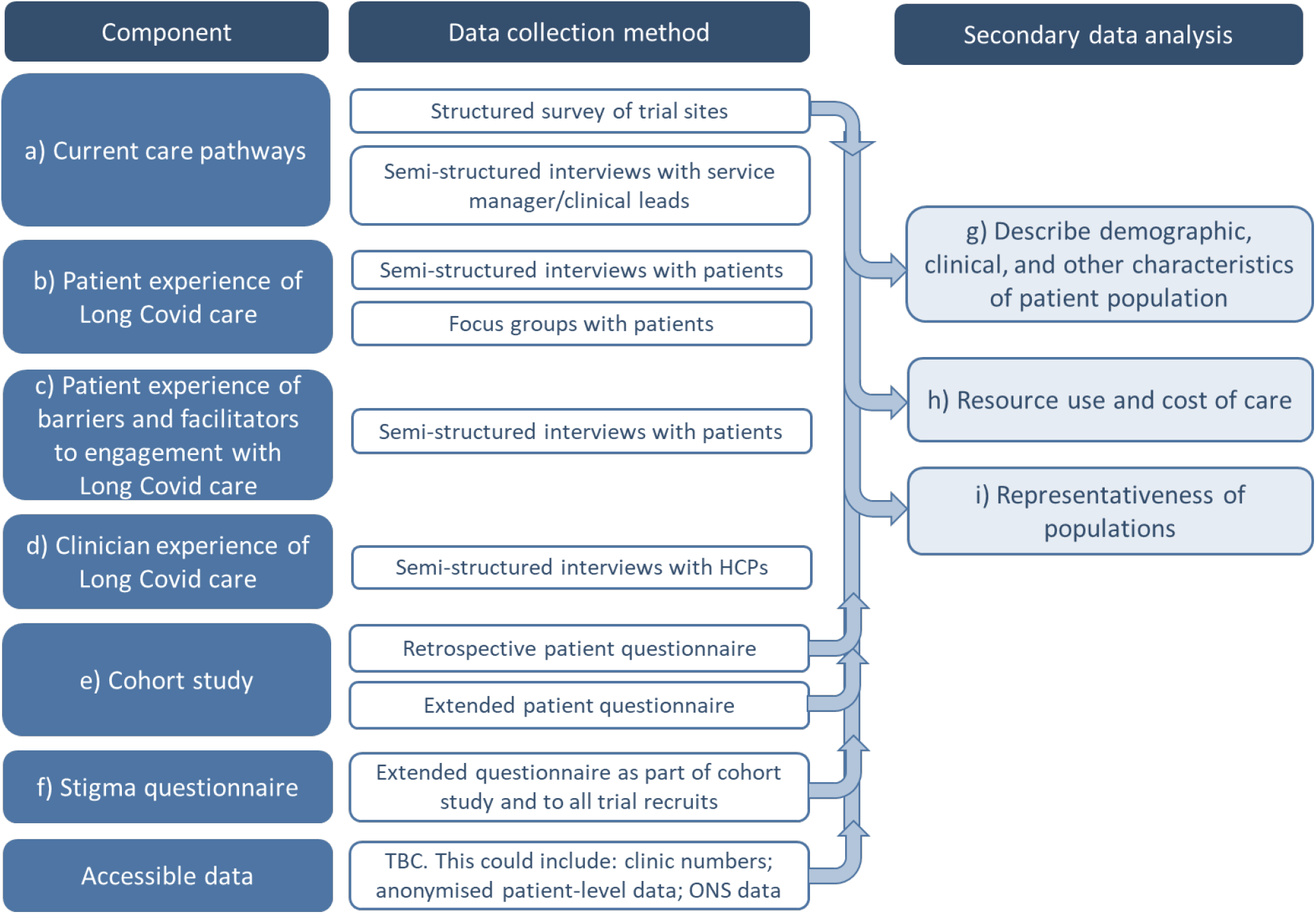
Study design for the CARE-INEQUAL work package for STIMULATE-ICP.

### Qualitative methods

#### Design

Qualitative methods will be used to understand current care pathways, patient and clinician experience of Long Covid services, and barriers and facilitators to care access. The four research components included are:

a. **Current care pathways at the 6 initial sites included in the STIMULATE-ICP clinical trial:** The service manager and/or clinical lead for each Long Covid clinic will be asked to complete a structured survey to collect key information related to the clinic set up and patient pathways. The survey will be followed with semi-structured interviews with the service manager and clinical lead to clarify any outstanding queries, and to understand their perception of the effectiveness of services and the sustainability of the clinics.
b. **Patient experience of current Long Covid care:** Both discharged and current patients will be invited to participate in either interviews (6 sites, n=4 at each site) or focus groups (1 at each of the 6 sites, n=6-8). They will be asked to describe their experience of care, including referral, assessment, diagnosis and treatment. This will provide an understanding of the patients’ perception of Long Covid care including any barriers to accessing care, their experience of the care pathway and any outcomes they experienced as a result of treatment.
c. **Patient experience of engagement with Long Covid care and stigma**: Semi-structured interviews will be carried out with Long Covid patients who have been referred to or accessed NHS Long Covid services (n=25-50). Patients will be asked to share their experiences of barriers and facilitators when accessing care. Interviews with people living with Long Covid but not accessing care will take place in a later phase of the study. This will be set out in a separate future protocol.
d. **Clinician experience of current Long Covid care:** Semi-structured interviews will be undertaken with health care professionals working in Long Covid clinics (6 sites, n=4 at each site), to explore their experiences of joining and working within the clinic, and their perception of inequalities in access to care for Long Covid patients. Participants will be recruited from four professional groups nominated by each site, to reflect the plurality of service design.

#### Data analysis

Interviews and focus groups will be carried out remotely using an online platform and will be recorded and stored securely. The data will be transcribed using a GDPR compliant transcribing company and will be anonymised and checked for accuracy. The data will be subject to thematic analysis, following Braun and Clarke’s thematic analysis framework [14] and using NVivo12 software.

### Cohort study and analysis of accessible data

#### Design

To explore the effectiveness of current Long Covid care, we will recruit to a cohort study those patients from sites which collected health outcome measures at the initial assessment as part of routine care. Patients at different points in their Long Covid pathway will be invited to complete health and wellbeing questionnaires they completed earlier in their care pathways.

e **Cohort study - existing data and extended questionnaire**: Patients at a minimum of one site (confirmed as University College London Hospitals NHS Trust, UCLH, and further sites will be identified in the initial site survey detailed in part (a) above) will be invited to participate in a cohort study, including analysis of their retrospective data, and completion of an extended questionnaire. At their initial clinic assessment, patients will be asked to complete a survey of their symptom nature and duration, and outcome measures. This study will aim to collect follow-up data on these measures to evaluate the extent of recovery in clinical and quality of life outcomes including: symptoms and employment status will be documented, and health and symptoms explored though the EQ-5D-5L questionnaire, Fatigue Assessment Scale (FAS), 2-item Patient Health Questionnaire (PHQ-2), and 2-item Generalised Anxiety Disorder (GAD-2) scale. Some additional questions will be asked in the follow-up questionnaire, including additional symptoms and whether symptoms have resolved, as well as asking patients to complete the Work and Social Adjustment Scale (WSAS) and the Perceived Deficits Questionnaire (PDQ5) (outcome measures included in the STIMULATE-ICP RCT).
g **Assessment of Long Covid stigma:** No validated Long Covid stigma scales currently exist. The one used for this study has been developed based on health stigma theory and emerging qualitative evidence of Long Covid stigma. It utilises items from existing scales related to stigmatising conditions (myalgic encephalomyelitis (ME)/chronic fatigue syndrome (CFS), depression and HIV). These items were reviewed by people living with Long Covid both internal (our patient and public involvement and engagement (PPIE) collaborators) and external to this study. As part of the Long Covid follow-up questionnaire in part (e), patients will be asked to complete a stigma questionnaire. This will be analysed along with other routine data to determine the prevalence and correlates of stigma and discrimination.

Additional data will be requested from the clinics, NHS England, NHS Digital and the Office for National Statistics (ONS) in order to:

g **Describe the Long Covid patient population:** Accessible patient data, such as those from the primary care electronic health record, will be analysed to understand the patient population in terms of age, gender, ethnicity, sociodemographic status, health care resource use, clinical and health outcomes.
h **Estimate the resource use and cost of care:** Patient reported data on resource use, costs and time of work, will be valued using published NHS costs (National Reference Costs; PSSRU Unit Costs of Health and Social Care) and other standard reference costs to estimate the overall societal costs of Long Covid care. Additionally, we will use clinic data collected in part (a) to calculate the costs of delivering care in Long Covid clinics and analyse how this varies by clinic and patient demographics.
i **Examine inequalities in Long Covid care:** Routinely collected socio-economic, demographic and health characteristics of patients accessing services will be compared to Office for National Statistics (ONS) data on COVID-19 infections and Long Covid in the community, to estimate representativeness of those reaching Long Covid services.

#### Data analysis – components (e), (g) and (h)

We will use statistical analysis and health economic methods to understand the associations between different symptoms, risk factors and Long Covid outcomes. We will investigate frequencies and the predictive value of different investigations and procedures employed in the assessment of patients referred to Long Covid clinics. We will study community, primary and secondary care healthcare utilisation in individuals with Long Covid at the different stages of diagnosis, treatment and management. We will also consider any confounding effect of comorbidities and multimorbidity by including them as covariates in regression analyses.

Data will be analysed to evaluate the extent of improvement in symptoms and health outcomes over time using appropriate methods, including t-tests and ANOVA. This will include, specifically, longitudinal change in EQ-5D-5L (including the accompanying visual analogue scale, EQ-VAS), employment status, FAS, PHQ-2, GAD-2 scales and resolution of previous symptoms. An evaluation of the extent of healthcare utilisation through the care pathway will be made.

Published costs will be applied to resource use to estimate the cost of Long Covid care. Combining the information on the Long Covid clinic provision with what it costs to deliver care at a patient level and estimates of the number of patients who will need Long Covid care, we will calculate the total cost of delivering care in Long Covid clinics in England over one year. In addition, we will describe the distribution of costs across primary and secondary care and the proportion of cost associated with diagnosis, pharmacological and non-pharmacological treatments including physical rehabilitation and counselling.

#### Data analysis – components (f) and (i)

Demographic and socioeconomic patient data as recorded in or obtained from Long Covid clinics (where available) will be analysed. They will be compared against demographic and socioeconomic characteristics of those with SARS-CoV-2 infection and self-reported Long Covid using the ONS COVID-19 Infection Survey (CIS) data. CIS is a representative national study of infection prevalence.

Where available, we will use the following variables to assess the representativeness of those accessing Long Covid services: age, gender, employment status, occupation, educational attainment, ethnicity, disability status, area-level index of Multiple Deprivation (full and income-domain), general health pre-COVID-19, pre-existing long term medical conditions including mental health conditions, lone parenting, regular care role, household size, insecure/temporary accommodation. Demographic and socioeconomic predictors of clinical outcomes as collected by Long Covid services will be explored.

We will also assess stigma prevalence and correlates using a quantitative Long Covid stigma short questionnaire. This scale is currently being validated in another study (not published-a follow up of a Long Covid community survey [9]). Demographic and socioeconomic correlates using the above variables where available will be assessed to identify characteristics most at risk of stigma. This will include descriptive statistics (means, medians, proportions); univariate comparisons (chi-square test for categorical variables and t-test for continuous variables); multivariable linear and logistic regression modelling.

### Patient and public involvement

STIMULATE-ICP has been enriched by robust patient and public involvement and engagement (PPIE). Each of our work packages has a PPIE co-lead, alongside clinical and academic co-leads, and PPIE features in our regular management and oversight meetings. In the work packages set out in this protocol, PPIE members have contributed to the research components, including informing the questionnaire design and approach to interviews, and will continue to be involved in the analysis plan and dissemination of findings.

### Ethics

Ethical approval for the project has been granted by the Berkshire Research Ethics Committee and the NHS HRA, REC reference: 22/SC/0047.

We will obtain written consent from participants for interviews and focus groups. Consent will be sought for prospective and retrospective data collection as part of the cohort study. Electronic consent will be used for the questionnaire and data access.

### Dissemination and outputs

The final dissemination plan is to be decided by the PPIE group members and the study Co-Is. However, dissemination will include publications, conference presentations, engagement with media and updates/publication on our study website (https://www.stimulate-icp.org/). Research participants will receive publications and reports generated by the study if they wish and where possible. In addition, we will disseminate via PPIE meetings during and at the end of the project.

## Discussion

As Long Covid is a new and emerging disease with a growing prevalence, it is paramount not only that efficacious treatments are developed, but that we also better understand the patient population, current care pathways, and the determinants of access to care, and thus ultimately inform effective guideline development. STIMULATE-ICP-CAREINEQUAL aims to address this evidence gap by describing current Long Covid care in the six initial clinics participating in the STIMULATE-ICP study and to understand potential barriers and facilitators to access to care.

There are currently fewer than 90 Long Covid clinics in England, with a variety of configurations according to local expertise. Rapid guidelines were produced in late 2020, with further policy published in June 2021, and funding to expand services beyond assessment to diagnostics and treatment. [15] However, many individuals seem to have been unable to access services, with long waiting lists [16] and temporary suspension of clinics in some areas over the 21/22 winter period contributing to this. [17] By undertaking this research, we will describe patient and clinician experiences of real-world barriers and facilitators to delivery and access of Long Covid services in a variety of service designs.

The strengths of this study include an inter-disciplinary approach, with patients, clinicians and qualitative and quantitative researchers involvement in the study design and execution. The six sites participating in the STIMULATE-ICP trial demonstrate a variety of service settings, design and duration; representing the diversity of Long Covid services in England. However, this may also be a challenge – both to the recruitment of participants, and to accessing baseline and quality patient data for our proposed analyses.

This study will provide insights into the deliverability of holistic Long Covid services; the impact of such services on health and care practitioners and Long Covid patients; the cost; and inequities in access. Further research is being undertaken within the broader project in order to make recommendations on improving access to Long Covid services. The data collected within this protocol will enrich the findings of the STIMULATE-ICP trial, which will explore the benefits of an integrated care pathway (ICP) for Long Covid and inform the STIMULATE-ICP-DELPHI sub-study [18], by providing a context for the implementation of any recommendations.

## Data Availability

This is a protocol paper; no data are available from this work at present.

## Authors Contributions

Study concepts and methodology were designed by MR, MG, PL, NAA, DC, MH, MP and AB. MR, PL, NAA, YM and DC wrote the original draft. Additional review and editing was carried out by GYHL, CvdF-C, DW, NW, HM, RMC, MG and EA.

## Acknowledgements

HM is supported by the National Institute for Health Research’s (NIHR’s) Comprehensive Biomedical Research Centre (BRC) at University College London Hospitals (UCLH).

NAA has lived experience of Long Covid and has contributed in an advisory role to World Health Organisation post-COVID-19 condition (PCC) definition and outcomes meetings and EU Commission’s Expert Panel on effective ways of investing in health in relation to PCC.

AB is supported by research funding from NIHR (including as PI of the STIMULATE-ICP study), European Union (part of the BigData@Heart Consortium, funded by the Innovative Medicines Initiative-2 Joint Undertaking under grant agreement No. 116074), British Medical Association (TP Gunton award), AstraZeneca and UK Research and Innovation.

MG is supported by the National Institute for Health and Social Care Research Applied Research Collaboration North West Coast (ARC NWC).

This paper has been published on behalf of the STIMULATE-ICP Consortium. An up-to-date version of Consortium members can be found: https://www.stimulate-icp.org/team

## Disclaimer

The views expressed in this publication are those of the author(s) and not necessarily those of the National Institute for Health and Socia Care Research or the Department for Health and Social Care.

